# Lower infant mortality and access to contraception reduce fertility in low- and middle-income nations

**DOI:** 10.1101/2021.12.16.21267946

**Authors:** Corey J A Bradshaw, Claire Perry, Chitra Maharani Saraswati, Melinda Judge, Jane Heyworth, Peter N Le Souëf

**Author notes:** **Correspondence to:** Peter N. Le Souëf, School of Medicine, The University of Western Australia, M431, 35 Stirling Hwy, Crawley, Western Australia 6009, Australia. These authors contributed equally to this work (Corey J A Bradshaw, Claire Perry, Peter N Le Souëf).

## Abstract

Although average contraceptive use has increased globally in recent decades, an estimated 222 million (26%) of women of child-bearing age worldwide face an unmet need for family planning — defined as a discrepancy between fertility preferences and contraception practice, or failing to translate desires to avoid pregnancy into preventative behaviours and practices.

While many studies have reported relationships between availability of contraception, infant mortality, and fertility, these relationships have not been evaluated quantitatively across a broad range of low- and middle-income countries. Using publicly available data from 46 low- and middle-income countries, we collated test and control variables in six themes: (*i*) availability of family planning, (*ii*) quality of family planning, (*iii*) maternal education, (*iv*) religion, (*v*) mortality, and (*vi*) socio-economic conditions. We predicted that higher nation-level availability/quality of family-planning services, maternal education, and wealth reduce average fertility, whereas higher infant mortality and religious adherence increase it. Given the sample size, we first constructed general linear models to test for relationships between fertility and the variables from each theme, from which we retained those with the highest explanatory power within a final general linear model set to determine the partial correlation of dominant test variables. We also applied boosted regression trees, generalised least-squares models, and a generalised linear mixed-effects models to account for non-linearity and spatial autocorrelation. On average among all countries, we found an association between all main variables and fertility, with reduced infant mortality having the strongest relationship with reduced fertility. Access to contraception was the next-highest correlate with reduced fertility, with female secondary education, home health visitations, and adherence to Catholicism having weak, if any, explanatory power. Our models suggest that decreasing infant mortality and increasing access to contraception will have the greatest effect on decreasing global fertility. We thus provide new evidence that progressing the United Nation’s Sustainable Development Goals for reducing infant mortality can be accelerated by increasing access to any form of family planning.

## INTRODUCTION

Although average contraceptive use has increased globally in recent decades, an estimated 222 million (26%) of women of child-bearing age worldwide face an unmet need for family planning — defined as a discrepancy between fertility preferences and contraception practice, or failing to translate desires to avoid pregnancy into preventative behaviours and practices^1^. The United Nation’s Sustainable Development Goals 3 and 5 emphasise the basic right to exercise control over sexual and reproductive health through universal access to family planning.^2^ While achieving Goal 3 is targeted for 2030, reducing global maternal mortality to < 70 per 100,000 live births and the under-5 mortality to ≤ 25 per 1,000 live births are not on track to be met.^3^ Providing readily available, high-quality family planning is necessary because this is expected to decrease not only fertility, but also the number of unintended pregnancies and infant and maternal deaths.^4 5^ Allowing individuals to be able to decide to have fewer children also has the potential to facilitate better investment in the overall health and well-being of families and communities.^5^

To date, there is no uniform measure (i.e., set of indicators) for availability and quality of family planning; therefore, gauging the effects these might have on fertility is difficult. Few studies have investigated the relationship between socio-economic conditions and fertility among nations; there are also few studies on the availability and quality of family planning that do more than just suggest a generally negative association with fertility.^6 7^ We investigated the association between fertility and the availability and quality of family planning, as well as the potential effects of education, religion, infant mortality, and socio-economic conditions. Specifically, we tested whether: (1) increasing the availability of family planning is associated with reducing fertility; (2) increasing the quality of family-planning services is associated with reducing fertility; (3) increasing female educational completion at primary and/or secondary school level is associated with reducing fertility; (4) increased fertility is observed in countries with a higher prevalence of religion that is against the use of artificial contraception; and (5) low socio-economic conditions and higher mortality are associated with higher fertility.

## METHODS

We used aggregated data at the national level, collating publicly available data from the Demographic and Health Surveys,^8^ Family Planning Effort Index,^9^ Multiple Indicator Cluster Surveys,^10^ National Composite Index on Family Planning,^11^ the World Bank,^12^ the World Factbook,^13^ and the World Health Organization Global Health Observatory data repositories.^14^ We obtained most of the required data from the Demographic and Health Surveys; these nationally representative surveys had uniform methods and included a similar period of reference when measuring indicators.

### Study populations

We used Demographic and Health Surveys datasets collected between 2010 to 2018 that contained data for all the required indicators, resulting in a selection of 46 countries (details in supplemental material Appendix 1). We aggregated these countries into the four regions of: Sub-Saharan Africa, South Asia/Pacific, Europe/Central Asia/Middle East/North Africa, and Latin America/Caribbean to account partially for spatial autocorrelation (see supplemental material Appendix 1, figure 1 and *Analyses*).

### Data

We derived the following test variables for model construction (additional information in Appendix 2). As the response, we used *fertility*, which is the mean number of children a woman has between the ages of 15 to 49 years,^15^ which we sourced from the World Bank (mean from 2010–2018).^12^ We broke down the modelling into to two phases (see *Analyses*) that first tested relationships to fertility within six separate themes: (***i***) availability of family planning, (***ii***) quality of family planning, (***iii***) maternal education, (***iv***) religion, (***v***) mortality, and (***vi***) socio-economic conditions. The second phase incorporated the top-ranked indicators into a final model set.

For **theme *i***, *availability of family-planning services* is measured via the ‘access’ index from the Family Planning Effort Index (2017 version).^9^ We added other indicators from the Demographic and Health Surveys and Multiple Indicator Cluster Surveys^10^ (most recent data per country) to measure other aspects of availability of family-planning not included in the ‘access’ index of the Family Planning Effort database,^9^ such as variable access to community health workers (extent of population visited by healthcare workers who educate about family planning and maternal and child health). For **theme *ii***, we derived *quality of family-planning services* using the ‘quality’ index from the National Composite Index on Family Planning^11^ (2017 version). We also included additional indicators from the Demographic and Health Surveys and Multiple Indicator Cluster Surveys^10^ (most recent data per country) to measure other aspects not included in the ‘quality’ index from the National Composite Index on Family Planning database.^11^ For **theme *iii***, we obtained *education* indicators using the following indicators from the Demographic and Health Surveys and Multiple Indicator Cluster Surveys^10^ (most recent data): (*i*) percentage of female primary school completion (for all countries), (*ii*) percentage of female secondary school completion (for all countries), (*iii*) percentage of male primary school completion (for 40 countries), and (*iv*) percentage of male secondary school completion (for 39 countries). We collected male education data to assess if there were major differences in the relationship to the availability and quality of family planning and/or fertility compared to female education. For **theme *iv*** (*religion*), we only included Catholicism in our analyses because this is the only religion with explicitly stated rules against artificial contraceptive use or family planning.^16 17^ Using the World Factbook^13^ (most recent data per country), we determined the percentage of a population who identified as Catholic for all countries. For **theme *v***, infant mortality is higher in areas of low socio-economics^18 19^ and a known correlate with fertility,^20^ so we included the most recent infant mortality data (deaths per 1000 live births) for each country from the Demographic and Health Surveys and Multiple Indicator Cluster Surveys.^10^ We also included the number of maternal deaths per 100,000 live births, and the number of conflict-related deaths per capita (most recent data from the World Bank^12^). For **theme *vi*** (*socio-economics*), women in the lowest 20% of household wealth have less availability of family planning compared with women from higher-wealth households.^21^ We therefore collated the following indicators as socio-economic measurements: (*a*) percentage of people residing in the lowest and highest wealth quintiles, (*b*) mean number of household members, and (*c*) percentage of households with three generations residing (from the Demographic and Health Surveys and Multiple Indicator Cluster Surveys^10^) (each country’s most recent data).

### Analyses

We applied descriptive analyses to provide an overview of the distribution across all variables. We first transformed all variables and scaled/centred them to improve homoscedasticity and Gaussian behaviour using a logit transformation of the base proportion and then centring and scaling using the *scale* function in the R programming language.^22^ We examined the transformed explanatory variables for collinearity using a non-parametric (Kendall’s *τ*) correlation matrix (supplemental table A1, appendix 3).

For the first modelling phase, we built general linear models with the *glm* function in R to identify the contributory (transformed) variable with the most explanatory power in each of the five themes (see supplemental appendices 4–7). We included various models in each theme and ranked them based on the Bayesian information criterion (BIC)^23 24^ given that our focus was on identifying the main drivers of variance in fertility as opposed to prediction,^25^ with relative model probability equal to its BIC weight (*w*BIC) (supplemental tables 2–5, appendices 4–7). Including the top-ranked variables from each theme into a final model set, we determined both the evidence for a non-random effect of the final variables on fertility, as well as the goodness of fit (percent deviance explained per model). We also built boosted-regression trees^26^ of the final model to account for potential nonlinearity in the relationships between the fertility response and the potential indicators (supplemental figures 2–4, appendices 4–7).

We suspected potential spatial autocorrelation among the country values, so we also constructed general linear mixed-effects models using the lme4 package^27^ in R, coding a random effect according to major world region (supplemental figure 1, appendix 1). Mixed-effects models potentially miss sub-regional spatial autocorrelation, so to account for a deeper level of spatial autocorrelation and to quantify uncertainty in the relationships between fertility and each explanatory variable, we resampled the dataset with replacement 1000 times. We then passed each resampled dataset to the boosted regression tree and then calculated the 2.5^th^ and 97.5^th^ percentiles for the respective distribution for each predicted fertility as the uncertainty bounds. We applied kappa (*κ*) limitation to the resampled selections to limit the influence of outliers,^28^ where we retained only the resampled mean ranks within *κσ* of the overall average mean (*κ* = 2). We then recalculated the average and standard deviation of the mean rank, with the process repeated five times.

Finally, we applied general least-squares models that are designed explicitly to account for spatial autocorrelation among spatial units (countries, in this case) to the final model set. For each country, we coded the centroid coordinates (in latitude/longitude) and determined that a spherical correlation was the top-ranked within-group correlation structure for the saturated model; we therefore ran the models in the final phase as per the general linear/mixed-effects models. We ranked the ensuing models according to *w*BIC, and calculated relative goodness-of-fit using three different pseudo-r^2^ metrics: McFadden, Cox and Snell, and Craig and Uhler (using the *nagelkerke* function in R library rcompanion^29^). All data and R code to repeat the analyses are provided at github.com/cjabradshaw/humanfertility.

## RESULTS

Fertility was Normally distributed (Shapiro-Wilk normality test: *W* = 0.973; *p* = 0.216) with one outlier (Niger), although that country’s value of 7.2 per woman is the highest national fertility globally.^12^ A non-parametric (Kendall’s *τ*) correlation matrix of the highest-ranked variable from each initial model (supplemental table 1, appendix 3) indicated that the strongest correlation observed was the relationship between access to contraception and infant mortality (*τ* = -0.447). Not included in supplemental table 1 are two relationships that suggested collinearity: female *versus* male primary education completion (*τ* = 0.9117) and female *versus* male secondary school completion (*τ* = 0.9534). We therefore removed male education both at the primary and secondary levels from further analysis. Of all regions, countries in Sub-Saharan Africa had the highest average number of infant mortalities (47 per 1000 live births), as well as the highest average fertility (4.9 per woman) (figure 1).

**Figure 1.**
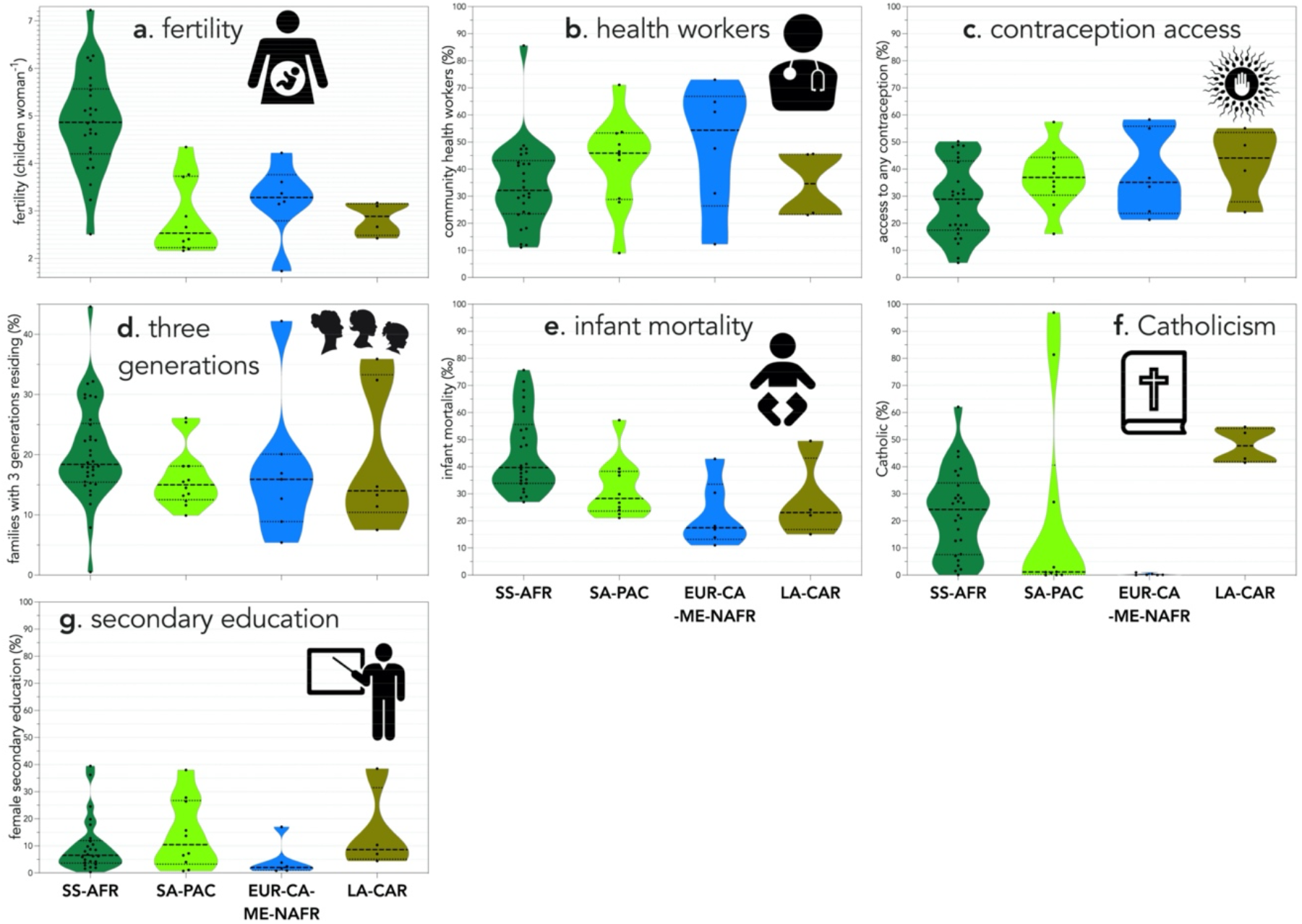
Violin plots of country-level raw values for **a**. fertility (children woman^-1^) and the highest-ranked variable from each of the six initial modelling phases — **b**. population with access to community health workers (%), **c**. population with access to any form of contraception (%), **d**. families with three generations residing in the same household (%), **e**. infant mortality (per 1000 births), **f**. population claiming adherence to Catholicism (%), and **g**. women who have achieved at least secondary school-level education (%),. The 46 low- and middle-income countries with full data included in the analysis are classed into four world regions (SS-AFR = Sub-Saharan Africa; SA-PAC = South Asia/Pacific; EUR-CA-ME-NAFR = Europe/Central Asia/Middle East/North Africa; LA-CAR = Latin America and Caribbean). Country codes per region (3-letter ISO): **SS-AFR** (BEN, BDI, CMR, TCD, COG, COD, GMB, GHA, KEN, LSO, LBR, MDG, MWI, MLI, MOZ, NAM, NER, NGA, RWA, SEN, ZAF, TZA, TGO, UGA, ZMB, ZWE); **SA-PAC** (BGD, KHM, IND, IDN, MMR, NPL, PAK, PNG, PHL, TLS), **EUR-CA-ME-NAFR** (ARM, EGY, JOR, KGZ, TJK, YEM); **LA-CAR** (DOM, GTM, HTI, HND)

### General linear / mixed-effects models

Access to community health workers was the most supported variable in phase 1 (access; supplemental table 2, supplemental figure 2, appendix 4), *access to any type of contraception* in phase 2 (quality; supplemental table A3, supplemental figure 3, appendix 5), *female secondary education completion* in phase 3 (education; supplemental table 4, appendix 6), *proportion Catholic* in phase 4 (religion), and *infant mortality* in phase 5 (socio-economics; supplemental table 5, supplemental figure 4, Appendix 7). Based on this group of five variables, we constructed 17 candidate general linear and general linear mixed-effects models (tables 2 and 3) to determine the most-supported models according to *w*BIC.

According to the final-phase general linear models, the model with access to community health workers and access to any form of contraception had the highest model support (*w*BIC = 69) and explained 58.9% of the deviance, followed by the saturated model (*w*BIC = 0.28; deviance explained = 66.4%) (Table 2). However, *infant mortality* had the highest % deviance explained (50.2%) among the single-variable models (Table 1).

**Table 1.** Candidate general linear models of the highest-ranked variable from each phase (1. availability, 2. quality, 3. education, 4. religion, 5. mortality, 6. socio-economics) in relation to variation in fertility among 46 low- and middle-income countries.

| model | *k*<sup>a</sup> | LL<sup>b</sup> | ΔBIC<sup>c</sup> | *wBIC*<sup>d</sup> | %DE<sup>e</sup> |
| --- | --- | --- | --- | --- | --- |
| CHW<sup>f</sup>+AC<sup>g</sup> | 3 | -66.26 | 0.00 | 0.69 | 58.9 |
| ALL<sup>h</sup> | 7 | -58.99 | 1.78 | 0.28 | 66.4 |
| CHW+IM<sup>i</sup> | 3 | -69.44 | 6.37 | 0.03 | 54.8 |
| AC+IM | 3 | -73.24 | 13.96 | <0.01 | 59.3 |
| IM+FSEC<sup>j</sup> | 3 | -77.47 | 22.42 | <0.01 | 51.5 |
| IM | 2 | -79.92 | 23.25 | <0.01 | 50.2 |
| CHW+FSEC | 3 | -78.27 | 24.03 | <0.01 | 29.7 |
| IM+GEN<sup>k</sup> | 3 | -78.34 | 24.18 | <0.01 | 52.8 |
| IM+CATH<sup>l</sup> | 3 | -78.95 | 25.39 | <0.01 | 50.4 |
| CHW+GEN | 3 | -80.05 | 27.58 | <0.01 | 31.0 |
| CHW | 2 | -82.21 | 27.84 | <0.01 | 24.7 |
| AC+CATH | 3 | -80.88 | 29.24 | <0.01 | 45.3 |
| CHW+CATH | 3 | -80.96 | 29.41 | <0.01 | 25.0 |
| AC+FSEC | 3 | -81.68 | 30.85 | <0.01 | 41.8 |
| AC | 2 | -83.88 | 31.17 | <0.01 | 41.2 |
| FSEC+GEN | 3 | -94.28 | 56.05 | <0.01 | 12.5 |
| FSEC | 2 | -97.13 | 57.67 | <0.01 | 3.3 |
| FSEC+CATH | 3 | -95.20 | 57.89 | <0.01 | 5.7 |
| GEN | 2 | -97.44 | 58.30 | <0.01 | 9.9 |
| CATH+GEN | 3 | -95.81 | 59.10 | <0.01 | 11.3 |
| intercept-only | 1 | -100.50 | 60.34 | <0.01 | 0.00 |
| CATH | 2 | -98.76 | 60.94 | <0.01 | 1.8 |

<sup>a</sup>*k* = number of parameters; <sup>b</sup>LL = log-likelihood; <sup>c</sup>ΔBIC = difference in Bayesian information criterion between model and top-ranked model; <sup>d</sup>*wBIC* = Bayesian information criterion weight ( ≈ model probability); <sup>e</sup>%DE = % deviance explained; <sup>f</sup>CHW = access to community health workers (variable of availability); <sup>g</sup>AC = access to any form of contraception; <sup>h</sup>ALL = saturated model (including the six highest-ranked variables, one from each phase); <sup>i</sup>IM = infant mortality; <sup>j</sup>FSEC = female secondary education completion; <sup>k</sup>GEN = proportion of households with three generations residing; <sup>l</sup>CATH = Catholicism.

For the generalised linear mixed-effects models accounting for the random effect of region, the saturated model was again ranked the highest (*w*AIC_*c*_ = 0.36), but the fixed effects accounted for only just over half of the total variance explained (*R*_m_ = 31.7% of 61.2%; Table 1). *Infant mortality* again had the highest explanatory power (*R*_m_= 27.6%) of any single-variable model (Table 2).

**Table 2.** Generalised linear mixed-effects models (only 10 top-ranked models according to *w*AIC_*c*_ shown) of the highest-ranked variable from each phase (1. availability, 2. quality, 3. education, 4. religion, 5. socio-economics) in relation to variation in fertility among 46 low- and middle-income countries.

| model | $k^a$ | LL<sup>b</sup> | $\Delta BIC^c$ | $wBIC^d$ | $R_m^e$ | $R_c^f$ |
| --- | --- | --- | --- | --- | --- | --- |
| CHW<sup>g</sup>+AC<sup>h</sup> | 5 | -59.02 | 0.00 | 0.88 | 27.6 | 63.5 |
| CHW+AC+IM<sup>i</sup> | 6 | -59.07 | 3.99 | 0.12 | 32.5 | 63.1 |
| CHW+IM | 5 | -67.09 | 16.23 | <0.01 | 24.3 | 50.8 |
| ALL<sup>j</sup> | 8 | -58.82 | 16.48 | <0.01 | 31.2 | 61.4 |
| AC+IM | 5 | -68.34 | 19.47 | <0.01 | 35.1 | 63.8 |
| AC | 4 | -71.09 | 20.91 | <0.01 | 19.4 | 62.5 |
| CHW+GEN<sup>k</sup> | 5 | -70.44 | 22.93 | <0.01 | 8.0 | 55.1 |
| CHW | 4 | -72.40 | 22.93 | <0.01 | 1.9 | 49.7 |
| CHW+FSEC<sup>l</sup> | 5 | -71.22 | 24.28 | <0.01 | 2.3 | 46.7 |
| AC+FSEC | 5 | -70.95 | 24.51 | <0.01 | 19.2 | 61.3 |
| CHW+CATH | 5 | -71.32 | 24.58 | <0.01 | 4.2 | 54.2 |
| AC+CATH<sup>m</sup> | 5 | -71.35 | 25.40 | <0.01 | 20.0 | 61.8 |
| ... |  |  |  |  |  |  |

<sup>a</sup> $k$ = number of parameters; <sup>b</sup>LL = log-likelihood; <sup>c</sup> $\Delta BIC$ = difference in Bayesian information criterion between model and top-ranked model; <sup>d</sup> $wBIC$ = Bayesian information criterion weight ( ≈ model probability); <sup>e</sup> $R_m$ = marginal $R^2$ ; <sup>f</sup> $R_c$ = conditional $R^2$ ; <sup>g</sup>CHW = access to community health workers; <sup>h</sup>AC = access to any form of contraception; <sup>i</sup>IM = infant mortality; <sup>j</sup>ALL = saturated model (including the six highest-ranked variables, one from each phase); <sup>k</sup>GEN = proportion of households with three generations residing; <sup>l</sup>FSEC = female secondary education completion; <sup>m</sup>CATH = Catholicism.

### Boosted regression trees

The boosted regression trees using either raw, untransformed values (grey bars in figure 2A), or the resampled relative contributions across 1000 iterations of the transformed variables (black bars in figure 2A), clearly indicated that *infant mortality* had the strongest explanatory power for variance in fertility among countries (figure 2A), followed by *access to any form of contraception* (figure 2A) (supplemental figures 2–4, appendices 4–8). The relationship between *infant mortality* and fertility was strongly stepped, suggesting a threshold of a sharp increase in fertility occurring once a country exceeded an infant mortality of ∼ 0.043 (figure 2B and 3). *Access to contraception* had a negative relationship with fertility, suggesting that fertility rates would decrease as a country improves its citizens’ access to contraception (figure 2C). While the relationships with the remaining three variables were in the hypothesised directions (figure 2D–F), their relative contributions were relatively weak (figure 2A).

**Figure 2.**
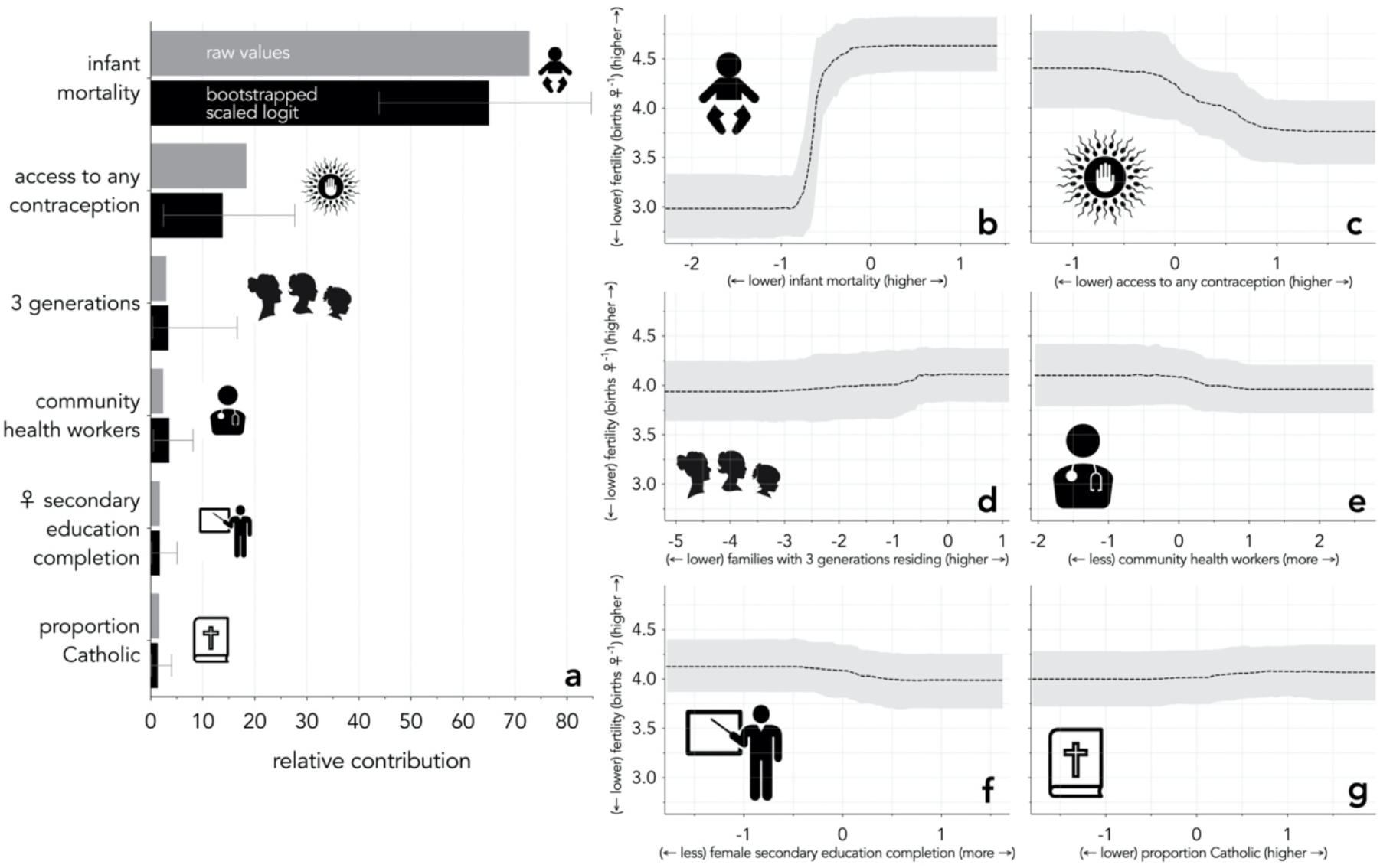
(**a**) The relative contribution and proportion of variance in indicators of availability, quality, education, religion, and socio-economics derived from boosted regression trees. The relative contribution of each variable (raw values) to the variance in fertility for each country in the dataset is represented by the grey bars. % deviance explained for raw and bootstrapped boosted regression trees were 42.7% and 37.2–38.3%, respectively. The black bars represent the resampled boosted regression trees for the same variables. Predicted fertility is expressed as a function of variation in (**b**) infant mortality, (**c**) access to any form of contraception, (**d**) proportion of families with three generations residing, (**e**) visits by a community health worker to discuss family-planning and maternal and child health, (**f**) female secondary education completion, and (**g**) proportion of Catholics.

**Figure 3.**
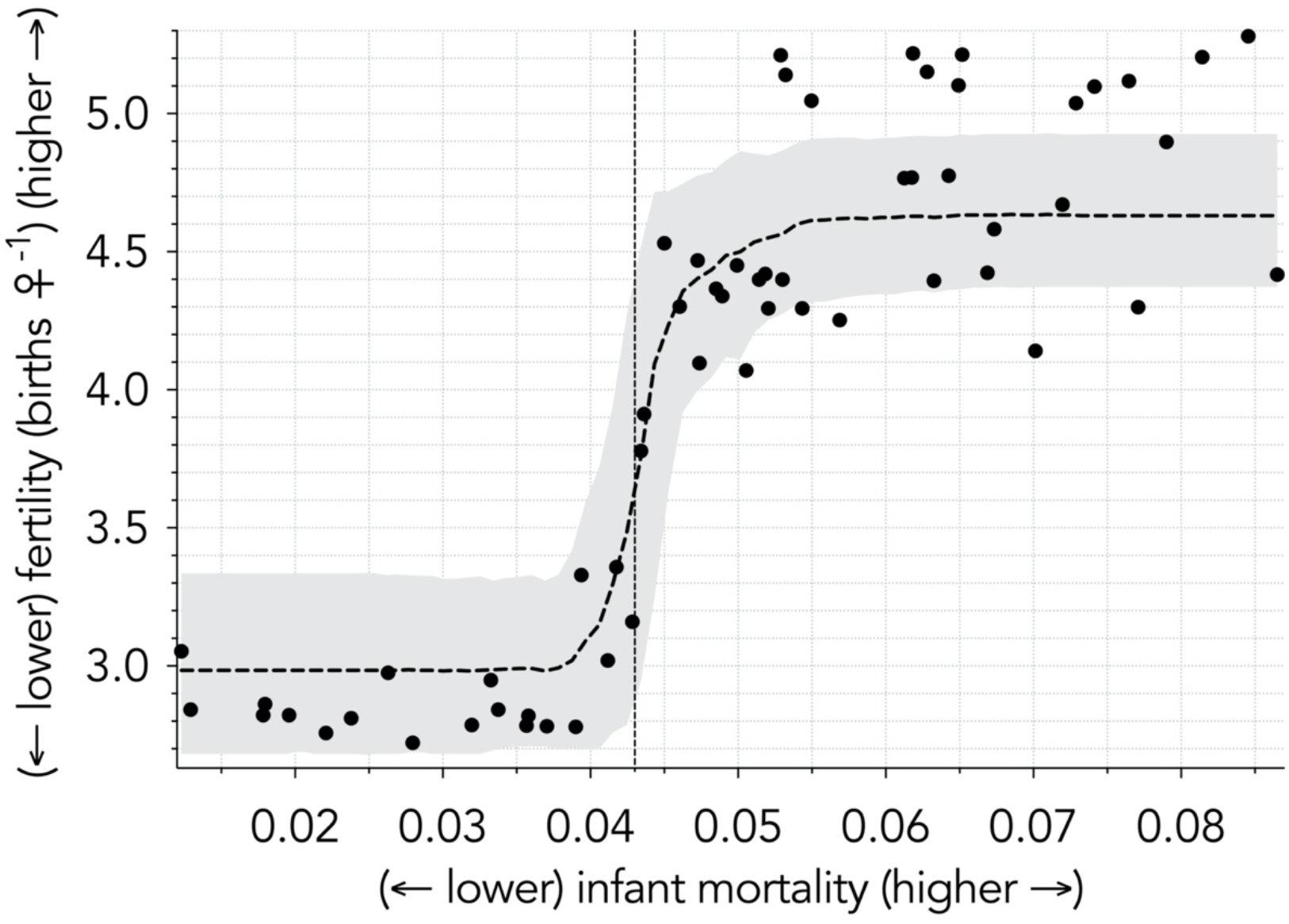
Predicted fertility as a function of variation in infant mortality (raw data from 59 countries for which both variables were available superimposed onto boosted regression tree relationship). Once infant mortality has exceeded a threshold of approximately 0.043 (43/1000 live births), fertility increases precipitously but then plateaus at 55/1000 live birth.

### General least-squares models

The country centroids explained 31.9–38.1% of the variation in the general least-squares models, and confirmed the dominance of both *infant mortality* and *access to any form of contraception* on fertility rates across countries (table 3).

**Table 3.** General least-squares models of the highest-ranked variable from each phase (*i*. availability, *ii*. quality, *iii*. education, *iv*. religion, *v*. mortality, *vi*. socio-economics) in relation to variation in fertility among 46 low- and middle-income countries.

| model | BIC<sup>a</sup> | *w*BIC<sup>b</sup> | psR<sup>2</sup>mcf<sup>c</sup> | psR<sup>2</sup>cs<sup>d</sup> | psR<sup>2</sup>cu<sup>e</sup> |
| --- | --- | --- | --- | --- | --- |
| AC<sup>f</sup> | 124.38 | 0.375 | 0.11 | 0.25 | 0.27 |
| AC+IM<sup>g</sup> | 125.67 | 0.197 | 0.13 | 0.29 | 0.31 |
| CHW<sup>h</sup>+AC | 125.78 | 0.131 | 0.13 | 0.29 | 0.31 |
| AC+FSEC<sup>i</sup> | 126.99 | 0.121 | 0.12 | 0.27 | 0.29 |
| IM | 127.98 | 0.080 | 0.08 | 0.19 | 0.20 |
| CHW+IM | 130.49 | 0.018 | 0.09 | 0.21 | 0.23 |
| IM+GEN<sup>j</sup> | 130.68 | 0.016 | 0.09 | 0.21 | 0.22 |
| IM+CATH<sup>k</sup> | 131.38 | 0.011 | 0.08 | 0.19 | 0.21 |
| IM+FSEC | 131.56 | 0.010 | 0.08 | 0.19 | 0.21 |
| CHW | 132.17 | 0.008 | 0.05 | 0.11 | 0.12 |
| *intercept-only* | 133.65 | 0.004 | <0.01 | <0.01 | <0.01 |
| CHW+GEN | 134.06 | 0.003 | 0.06 | 0.15 | 0.16 |
| GEN | 133.90 | 0.003 | 0.03 | 0.08 | 0.08 |
| ALL<sup>l</sup> | 137.52 | 0.001 | 0.16 | 0.34 | 0.37 |
| CHW+FSEC | 135.97 | 0.001 | 0.05 | 0.11 | 0.12 |
| CHW+CATH | 135.94 | 0.001 | 0.05 | 0.11 | 0.12 |
| FSEC | 137.48 | 0.001 | <0.01 | <0.01 | <0.01 |
| CATH | 137.47 | 0.001 | <0.01 | <0.01 | <0.01 |
| FSEC+CATH | 141.30 | <0.001 | <0.01 | <0.01 | <0.01 |
| FSEC+GEN | 137.72 | <0.001 | <0.01 | <0.01 | <0.01 |
| AC+CATH | 149.82 | <0.001 | -0.07 | -0.2 | -0.22 |
| CATH+GEN | 137.72 | <0.001 | 0.03 | 0.08 | 0.08 |

<sup>a</sup>BIC = Bayesian information criterion; <sup>b</sup>*w*BIC = Bayesian information criterion weight ( ≈ model probability); <sup>c</sup>psR<sup>2</sup>mcf = pseudo-R<sup>2</sup> (McFadden metric); <sup>d</sup>psR<sup>2</sup>cs = pseudo-R<sup>2</sup> (Cox & Snell metric); <sup>e</sup>psR<sup>2</sup>cu = pseudo-R<sup>2</sup> (Craig & Uhler metric); <sup>f</sup>AC = access to any form of contraception; <sup>g</sup>IM = infant mortality; <sup>h</sup>CHW = access to community health workers; <sup>i</sup>FSEC = female secondary education completion; <sup>j</sup>GEN = proportion of families with three generations residing; <sup>k</sup>CATH = Catholicism; <sup>l</sup>ALL = saturated model (including the six highest-ranked variables, one from each phase).

## DISCUSSION

Our study is the first to investigate the potential associations between the availability and quality of family planning and fertility, while simultaneously considering other potential contributory variables among 46 low- and middle-income countries. We found that high infant mortality was most strongly related to high fertility, but nonlinearly, followed by reduced access to contraception. However, the relative contribution of each of these variables to reducing fertility potentially also needs to consider the correlation between access to contraception and decreased infant mortality. Increased access to contraception has been previously associated with decreased infant mortality and is thought to act through increasing space between births, and avoiding the increased infant mortality found with higher birth order.^30 31^ We also found that other factors considered to be important in reducing fertility, including community health worker visits, female education, and religion (Catholicism) had only weak associations, at least at the spatial scale across nations.

Infant mortality as a strong predictor of variation in fertility is supported elsewhere.^20^ Overall, our findings support the notion that to decrease global fertility, both infant survival rates plus access to contraception need to be increased. Recommendations for measures to decrease infant mortality emphasise improving the quality of antenatal care, increasing the number of trained healthcare staff at births, and improving postnatal care for both infants and mothers.^32 33^ A greater emphasis on providing access to contraception as a direct contribution to decreasing infant mortality is important for such guidelines.

There are multiple factors that can influence fertility, some of the subtleties of which we could not test directly with the available data. Responsible family planning requires the effort of both men and women, yet male contraceptive use is decreasing globally.^15^ Potential reasons could be local views on gender equality, respect and dignity for women, and the proportion of women participating in the workforce.^34^ Child marriage and gender-based violence are positively correlated with low contraceptive use and increased fertility in some conditions;^35 36^ however, women who experience child-marriage have a higher modern contraceptive use (e.g., female and male sterilisation, intra-uterine device, oral hormonal pills, vaginal barrier methods) compared to adult married women.^37^ Our findings also suggest that increasing the availability of contraceptives can be effective in reducing fertility where parents do not have access to secondary education. Fundamentally, contraceptive use is closely linked to infant mortality. By allowing citizens to choose family planning by providing readily available, modern methods of contraception could improve infant survival because parents can plan and space their births, thereby investing in higher-quality care for each child. This could also lead to improvements in maternal and child-health outcomes globally.

Another strength of our study is using nationally representative surveys from the Demographic and Health Surveys database, which suggests general applicability to low- and middle-income nations. Further, applying several different modelling frameworks that confirmed the main contributors added robustness to our findings. Our design meant that possible biases exist, such as potentially hiding spatial variation within countries when relying on national averages potentially. There was also the potential for systematic differences between countries in terms of data collection and reporting (i.e., unstated variation in the number of surveys completed for each country).

Improvements need to continue to increase infant survival, particularly in low- and middle-income countries, which would in turn reduce fertility to and improve child and maternal health outcomes. An important component of these activities is in providing both women and men the choice to access non-coercive, quality family-planning services. Overall, there is more that can be done to aid in meeting the initiatives of the Sustainable Development Goals of the United Nations, which, if unmet, will see a global increase in fertility, more child deaths, and more birth-related deaths among women.

## Data Availability

All data and R code to repeat the analyses are provided at github.com/cjabradshaw/humanfertility

## Contributors

### Funding

We acknowledge financial support from the University of Western Australia.

### Patient consent for publication

Not applicable

### Ethics approval

Since all data used in this study were already in the public domain, ethical approval was not required.

### Provenance and peer review

Not commissioned; externally peer reviewed.

### Data availability statement

All data and R code to repeat the analyses are provided at github.com/cjabradshaw/humanfertility.

## Supplemental material

## Appendix 1 The four world regions

Given minimum sample-size requirements for each level of the random effect, we settled on four main regions that had sufficient replication: (1) South Asia / Pacific (*n* = 10), Europe / Central Asia / Middle East / North Africa (*n* = 6), sub-Saharan Africa (*n* = 26), and Latin America / Caribbean (*n* = 4) (supplemental figure 1).

**Figure 1.**
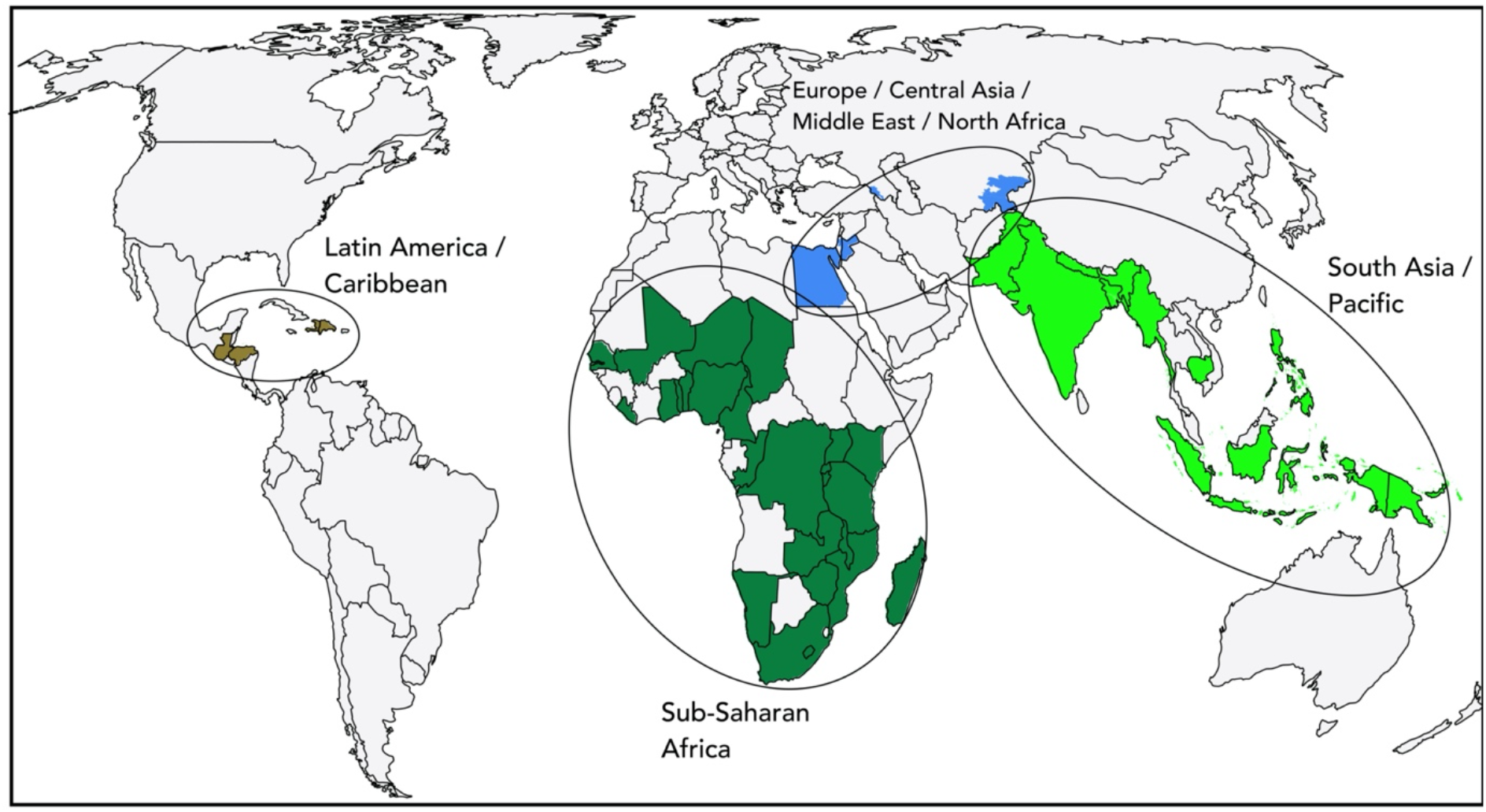
World map showing four regional classes used as a random effect in the general linear mixed-effects models (*n* = 46 countries). Country codes per region (3-letter ISO): **Sub-Saharan Africa** (BEN, BDI, CMR, TCD, COG, COD, GMB, GHA, KEN, LSO, LBR, MDG, MWI, MLI, MOZ, NAM, NER, NGA, RWA, SEN, ZAF, TZA, TGO, UGA, ZMB, ZWE); **South Asia/Pacific** (BGD, KHM, IND, IDN, MMR, NPL, PAK, PNG, PHL, TLS), **Europe/Central Asia/Middle East/North Africa** (ARM, EGY, JOR, KGZ, TJK, YEM); **Latin America/Caribbean** (DOM, GTM, HTI, HND)

## Appendix 2 Description of indicators within each index

Availability of family planning is based on the ‘access’ index in the Family Planning Effort database^9^: (*a*) access to the intrauterine device, (*b*) access to contraceptive pills, (*c*) access to injectables, (*d*) access to female sterilisation, (*e*) access to male sterilisation, (*f*) access to condoms, (*g*) access to implants, (*h*) access to emergency contraception, (*i*) access to safe abortion, (*j*) access to intrauterine device removal, (*k*) access to implant removal, and (*l*) sterilisation permanence explained by a trained healthcare worker.

Additional indices from the Demographic and Health Surveys^8^ and Multiple Indicator Cluster Surveys^10^ include: (*a*) involvement of private-sector agencies and groups, (*b*) community-based distribution, (*c*) social marketing (extent of coverage by a social marketing program that subsidise contraceptive sales), (*d*) community health workers (extent of population visited by healthcare workers to educate about family planning and maternal and child health), and (*e*) logistics and transport (extent to which logistics and transport networks are sufficient in maintaining contraceptive supplies and equipment at all services, time and levels).

The quality of the family planning index from the National Composite Index on Family Planning^11^ includes: (*a*) standard of practices in line with the World Health Organization, (*b*) guidelines on task-sharing, (*c*) indicators used by public family planning services, (*d*) indicators used by private family planning services, (*e*) structures in place to address quality, (*f*) information collected regarding informed choice and provider bias, (*g*) training programs for workers, (*h*) logistics and transport able to supply sufficient, quality services, (*i*) adequate supervision and monitoring in place, (*j*) sterilisation permanence education to clients, (*k*) proportion of population who have access to intrauterine device removal, (*l*) proportion of population who have access to implant removal. Additional indices from the Demographic and Health Surveys^8^ and Multiple Indicator Cluster Surveys^10^ include: (*a*) percentage of the population using any form of contraception, (*b*) percentage of the population using a modern contraceptive method, (*c*) percentage of the population using a traditional contraceptive method, and (*d*) percentage of a population using no form of contraceptive method.

## Appendix 3 Correlation among main explanatory variables

**Table 1.** Correlation (Kendall’s *τ*) matrix of the highest-ranked variable from each of the six thematic modelling phases (1. availability, 2. quality, 3. education, 4. religion, 5. mortality, 6. socio-economics) among 46 low- and middle-income countries.

|  | **access<sup>a</sup>** | **Cathol<sup>b</sup>** | **comm work<sup>c</sup>** | **infant mort<sup>d</sup>** | **secondary completion** |
| --- | --- | --- | --- | --- | --- |
| **Cathol** | -0.053 |  |  |  |  |
| **comm work** | 0.107 | -0.274 |  |  |  |
| **infant mort** | -0.447 | 0.134 | -0.293 |  |  |
| **secondary completion<sup>e</sup>** | -0.147 | 0.241 | -0.076 | 0.148 |  |
| **gen<sup>f</sup>** | -0.276 | 0.207 | -0.202 | 0.252 | 0.192 |

<sup>a</sup>**access** = access to any form of contraception; <sup>b</sup>**Cathol** = proportion of Catholics; <sup>c</sup>**comm work** = visitation by a community healthcare worker; <sup>d</sup>**infant mort** = infant mortality; <sup>e</sup>**secondary completion** = female secondary school completion; <sup>f</sup>**gen** = proportion of households with three generations residing

## Appendix 4 Availability of family planning

**Table 2.** General linear models for indicators of availability of family-planning in relation to variation in fertility among 50 low- and middle-income countries.

| model | *k*<sup>a</sup> | LL<sup>b</sup> | ΔBIC<sup>c</sup> | wBIC<sup>d</sup> | %DE<sup>e</sup> |
| --- | --- | --- | --- | --- | --- |
| ACC<sup>f</sup> | 2 | -81.66 | 0.00 | 0.18 | 26.4 |
| ACC+CBDT<sup>g</sup> | 3 | -79.96 | 0.67 | 0.13 | 31.2 |
| CHW<sup>h</sup> | 2 | -82.21 | 1.10 | 0.10 | 24.7 |
| ACC+SOCM<sup>i</sup> | 3 | -80.20 | 1.16 | 0.10 | 30.5 |
| ACC+LOGT<sup>j</sup> | 3 | -80.21 | 1.17 | 0.10 | 30.5 |
| SOCM+CHW | 3 | -80.56 | 1.88 | 0.07 | 29.5 |
| SOCM | 2 | -82.90 | 2.47 | 0.05 | 22.6 |
| CBDT+CHW | 3 | -78.95 | 2.56 | 0.05 | 28.6 |
| PRIV<sup>k</sup> | 2 | -80.90 | 3.30 | 0.03 | 21.3 |
| ACC+PRIV | 3 | -81.45 | 3.66 | 0.03 | 27.0 |
| LOGT | 2 | -83.52 | 3.71 | 0.03 | 20.7 |
| CBDT | 2 | -83.56 | 3.79 | 0.03 | 20.6 |
| ACC+CHW | 3 | -81.55 | 3.85 | 0.03 | 26.7 |
| CHW+LOGT | 3 | -81.79 | 4.33 | 0.02 | 26.0 |
| PRIV+CHW | 3 | -81.99 | 4.74 | 0.02 | 25.4 |
| SOCM+LOGT | 3 | -82.44 | 5.63 | 0.01 | 24.0 |
| CBDT+SOCM | 3 | -82.58 | 5.92 | <0.01 | 23.6 |
| PRIV+SOCM | 3 | -82.85 | 6.45 | <0.01 | 22.8 |
| ACC+CBDT+SOCM+LOGT | 5 | -79.15 | 7.21 | <0.01 | 33.4 |
| PRIV+LOGT | 3 | -83.26 | 7.28 | <0.01 | 21.5 |
| PRIV+CBDT | 3 | -83.30 | 7.35 | <0.01 | 21.4 |
| CBDT+LOGT | 3 | -83.51 | 7.77 | <0.01 | 20.7 |
| ACC+CBDT+SOCM+CHW+LOGT | 6 | -77.65 | 8.28 | <0.01 | 37.3 |
| ALL<sup>l</sup> | 7 | -77.64 | 12.34 | <0.01 | 37.3 |
| intercept-only | 1 | -100.5 | 33.61 | <0.01 | 0.0 |

<sup>a</sup>*k* = number of parameters; <sup>b</sup>LL = log-likelihood; <sup>c</sup>ΔBIC = difference in Bayesian information criterion between model and top-ranked model; <sup>d</sup>wBIC = Bayesian information criterion weight ( ≈ model probability); <sup>e</sup>%DE = % deviance explained; <sup>f</sup>ACC = ‘access’ index comprising indicators for availability of family planning from the Family Planning Effort Index<sup>9</sup>; <sup>g</sup>CBDT = community-based distribution of family planning; <sup>h</sup>CHW = visitation by a community health worker; <sup>i</sup>SOCM = social marketing of subsidised contraceptives; <sup>j</sup>LOGT = logistics and transport; <sup>k</sup>PRIV = involvement of private-sector agencies and groups; <sup>l</sup>ALL = saturated model (all variables included).

**Figure 2.**
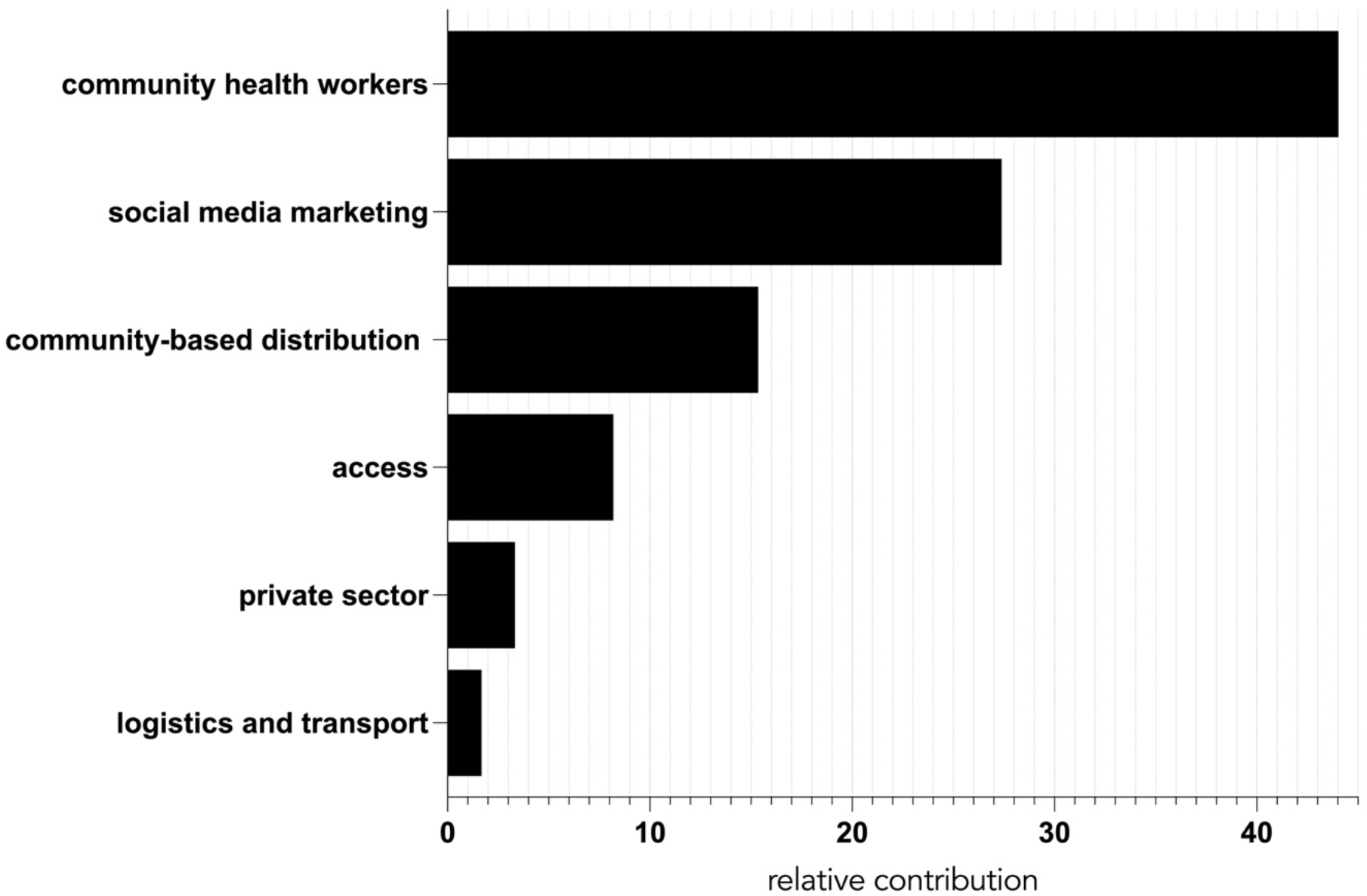
Boosted regression tree results (variable relative performance) for availability of family planning among 50 low- and middle-income countries. **community health workers** = visitation by a community health worker; **social media marketing** = social marketing of subsidised contraceptives; **community-based distribution** = community-based distribution of family-planning; **access** = ‘access’ index comprising of indicators for availability of family-planning from the Family Planning Effort Index^9^; **private sector** = involvement of private-sector agencies and groups; **logistics and transport** = logistics and transport;

## Appendix 5 Quality of family planning

**Table 3.** General linear models for indicators of quality of family-planning in relation to variation in fertility among 41 low- and middle-income countries.

| model | *k*<sup>a</sup> | LL<sup>b</sup> | ΔBIC<sup>c</sup> | wBIC<sup>d</sup> | %DE<sup>e</sup> |
| --- | --- | --- | --- | --- | --- |
| QUAL<sup>f</sup>+AC<sup>g</sup> | 3 | -71.14 | 0.00 | 0.43 | 49.8 |
| QUAL+NC<sup>h</sup> | 3 | -71.14 | 0.00 | 0.43 | 49.8 |
| QUAL+MC<sup>i</sup> | 3 | -72.90 | 3.53 | 0.07 | 46.1 |
| ALL<sup>j</sup> | 6 | -69.12 | 4.12 | 0.06 | 53.8 |
| QUAL+TC<sup>k</sup> | 3 | -78.12 | 13.97 | <0.01 | 33.2 |
| QUAL | 2 | -83.50 | 20.64 | <0.01 | 20.7 |
| TC+NC | 3 | -81.61 | 20.94 | <0.01 | 45.7 |
| AC+TC | 3 | -81.61 | 20.95 | <0.01 | 45.7 |
| NC | 2 | -83.88 | 21.40 | <0.01 | 41.2 |
| AC | 2 | -83.88 | 21.40 | <0.01 | 41.2 |
| MC+NC | 3 | -81.96 | 21.64 | <0.01 | 45.0 |
| AC+MC | 3 | -81.96 | 21.65 | <0.01 | 45.0 |
| AC+NC | 3 | -83.79 | 25.31 | <0.01 | 41.4 |
| MC+TC | 3 | -84.84 | 27.40 | <0.01 | 39.3 |
| MC | 2 | -92.28 | 38.20 | <0.01 | 21.5 |
| TC | 2 | -93.37 | 40.38 | <0.01 | 18.4 |
| intercept-only | 1 | -100.5 | 50.57 | <0.01 | 0.0 |

<sup>a</sup>*k* = number of parameters; <sup>b</sup>LL = log-likelihood; <sup>c</sup>ΔBIC = difference in Bayesian information criterion between model and top-ranked model; <sup>d</sup>wBIC = Bayesian information criterion weight ( ≈ model probability); <sup>e</sup>%DE = % deviance explained; <sup>f</sup>QUAL = 'quality' index from the National Composite Index of Family Planning<sup>11</sup>; <sup>g</sup>AC = access to any form of contraception; <sup>h</sup>NC = access to no form of contraception; <sup>i</sup>MC = access to modern contraceptives; <sup>j</sup>ALL = saturated model (all variables included); <sup>k</sup>TC = access to traditional contraceptives.

**Figure 3.**
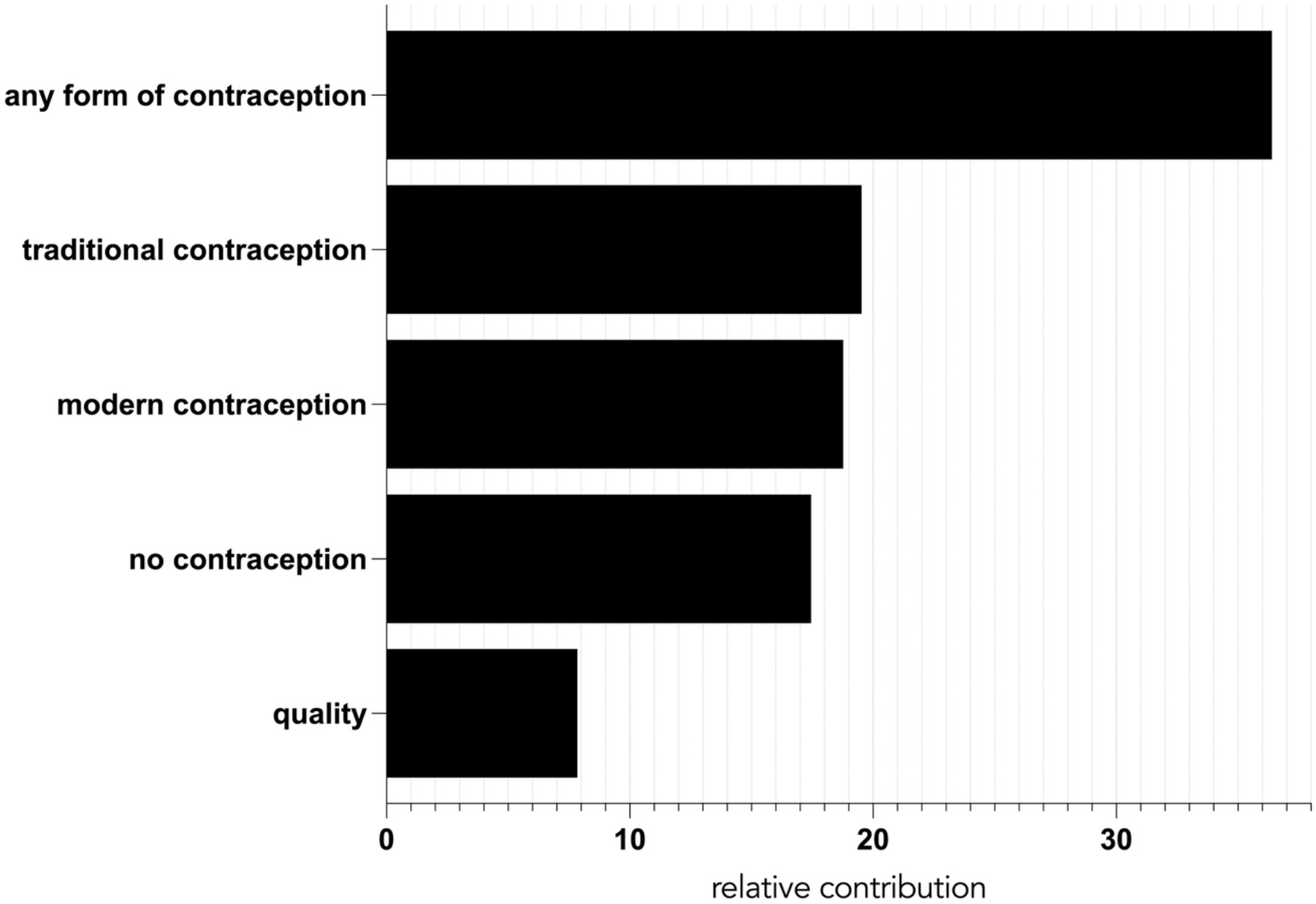
Boosted regression tree results (variable relative performance) for quality of family planning among 41 low- and middle-income countries. **any form of contraception =** proportion of a population who have access to any form (modern and/or traditional) contraception; **traditional contraception =** proportion of a population who have access to traditional forms of contraception only; **modern contraception =** proportion of a population who have access to modern forms of contraception; **no contraception =** proportion of a population who have no access to contraception; **quality =** ‘quality’ index of family-planning indicators from the National Composite Index on Family Planning^*11*^.

## Appendix 6 Education

**Table 4.** General linear models for indicators of education in relation to fertility among 57 low- and middle-income countries.

| model | $k^a$ | LL<sup>b</sup> | ΔBIC<sup>c</sup> | wBIC<sup>d</sup> | %DE<sup>e</sup> |
| --- | --- | --- | --- | --- | --- |
| **FSEC<sup>f</sup>** | 2 | -97.13 | 0.00 | 0.62 | 3.3 |
| *intercept-only* | 1 | -100.5 | 2.67 | 0.16 | 0.0 |
| **FPEC<sup>g</sup>** | 2 | -98.91 | 3.56 | 0.10 | 1.3 |
| **FPEC+FSEC** | 3 | -96.96 | 3.73 | 0.10 | 3.9 |
| **FPEC+FSEC+FPEC\*FSEC** | 4 | -96.70 | 7.30 | 0.02 | 4.7 |

<sup>a</sup> $k$ = number of parameters; <sup>b</sup>LL = log-likelihood; <sup>c</sup> $\Delta$ BIC = difference in Bayesian information criterion between model and top-ranked model; <sup>d</sup>wBIC = Bayesian information criterion weight ( ≈ model probability); <sup>e</sup>%DE = % deviance explained; <sup>f</sup>FSEC = female secondary education completion; <sup>g</sup>FPEC = female primary education completion.

## Appendix 7 Mortality

**Table 5.** General linear models for indicators of mortality in relation to fertility among 55 low- and middle-income countries.

| model | $k^a$ | LL<sup>b</sup> | $\Delta\text{BIC}^c$ | $w\text{BIC}^d$ | %DE<sup>e</sup> |
| --- | --- | --- | --- | --- | --- |
| **IM<sup>f</sup>+CRD<sup>g</sup>** | 3 | -76.56 | 0.00 | 0.37 | 53.0 |
| **IM+MM<sup>h</sup>+CRD** | 4 | -74.77 | 0.49 | 0.29 | 53.2 |
| **IM** | 2 | -79.92 | 2.65 | 0.10 | 50.2 |
| **IM+MM** | 3 | -78.19 | 3.25 | 0.07 | 50.3 |
| **IM+CRD+IM\*CRD** | 4 | -76.47 | 3.89 | 0.05 | 53.2 |
| **IM+MM+CRD+IF\*MM** | 5 | -74.58 | 4.19 | 0.05 | 53.5 |
| **IM+MM+CRD+IF\*CRD** | 5 | -74.65 | 4.34 | 0.04 | 53.4 |
| **IM+MM+IF\*MM** | 4 | -78.07 | 7.09 | 0.01 | 50.5 |
| **IM+MM+CRD+IF\*MM+IF\*CRD** | 6 | -74.48 | 8.08 | 0.01 | 53.6 |
| **MM+CRD** | 3 | -93.68 | 34.23 | <0.01 | 6.8 |
| **CRD** | 3 | -97.03 | 36.87 | <0.01 | 3.6 |
| **MM** | 2 | -97.12 | 37.04 | <0.01 | 3.3 |
| **intercept-only** | 1 | -100.50 | 39.73 | <0.01 | 0.0 |

<sup>a</sup> $k$ = number of parameters; <sup>b</sup>LL = log-likelihood; <sup>c</sup> $\Delta\text{BIC}$ = difference in Bayesian information criterion between model and top-ranked model; <sup>d</sup> $w\text{BIC}$ = Bayesian information criterion weight ( ≈ model probability); <sup>e</sup>%DE = % deviance explained; <sup>f</sup>IM = infant mortality (deaths per 1000 births); <sup>g</sup>CRD = conflict-related deaths per capita; <sup>h</sup>MM = maternal mortality (deaths per 100,000 live births).

**Figure 4.**
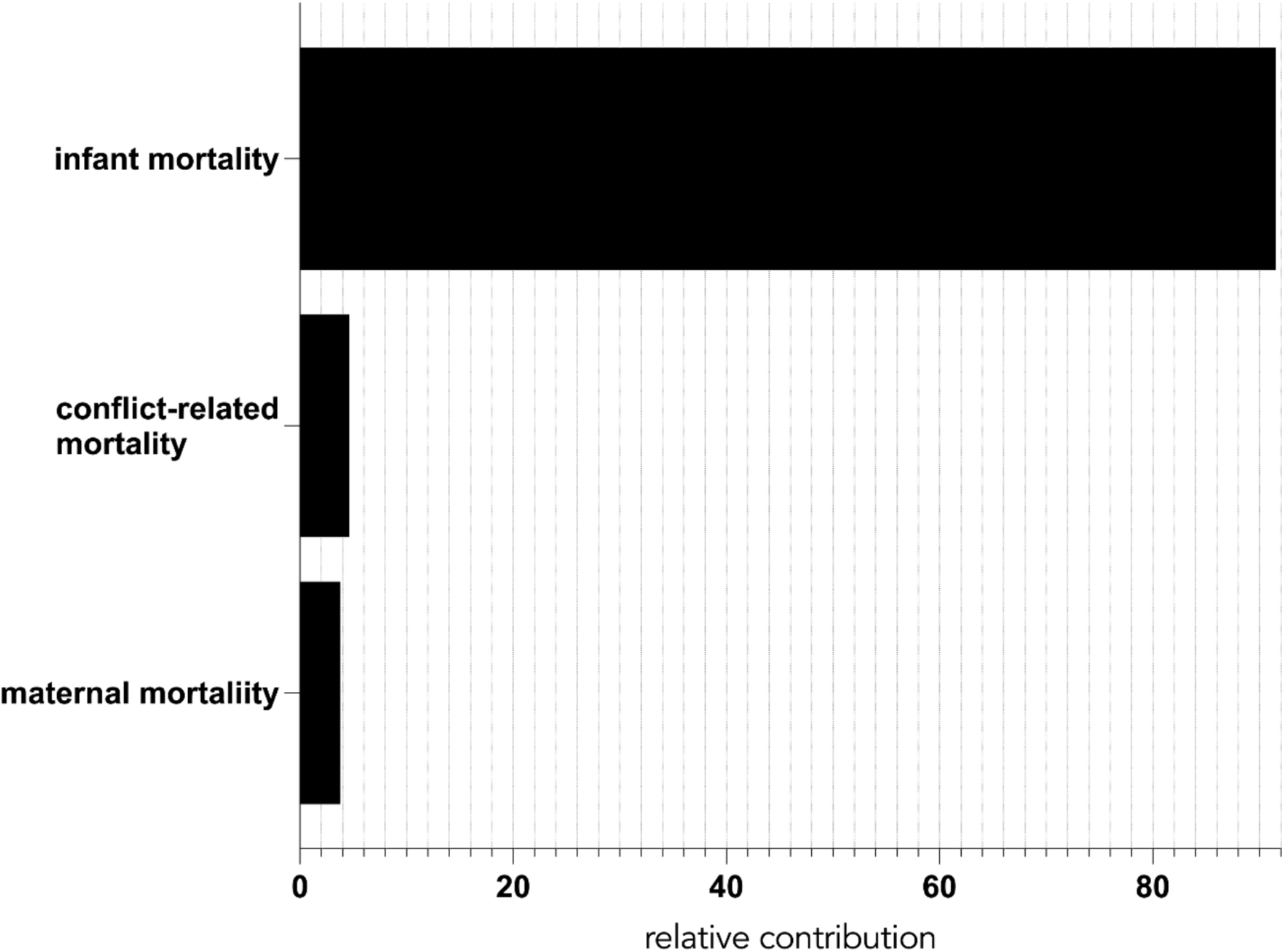
Boosted regression tree results (variable relative performance) for mortality among 55 low- and middle-income countries. **infant mortality** (deaths per 1000 births); **conflict-related mortality** (deaths per capita); **maternal mortality** (deaths per 100,000 live births)

## Appendix 8 Socio-economics

**Table 6.**
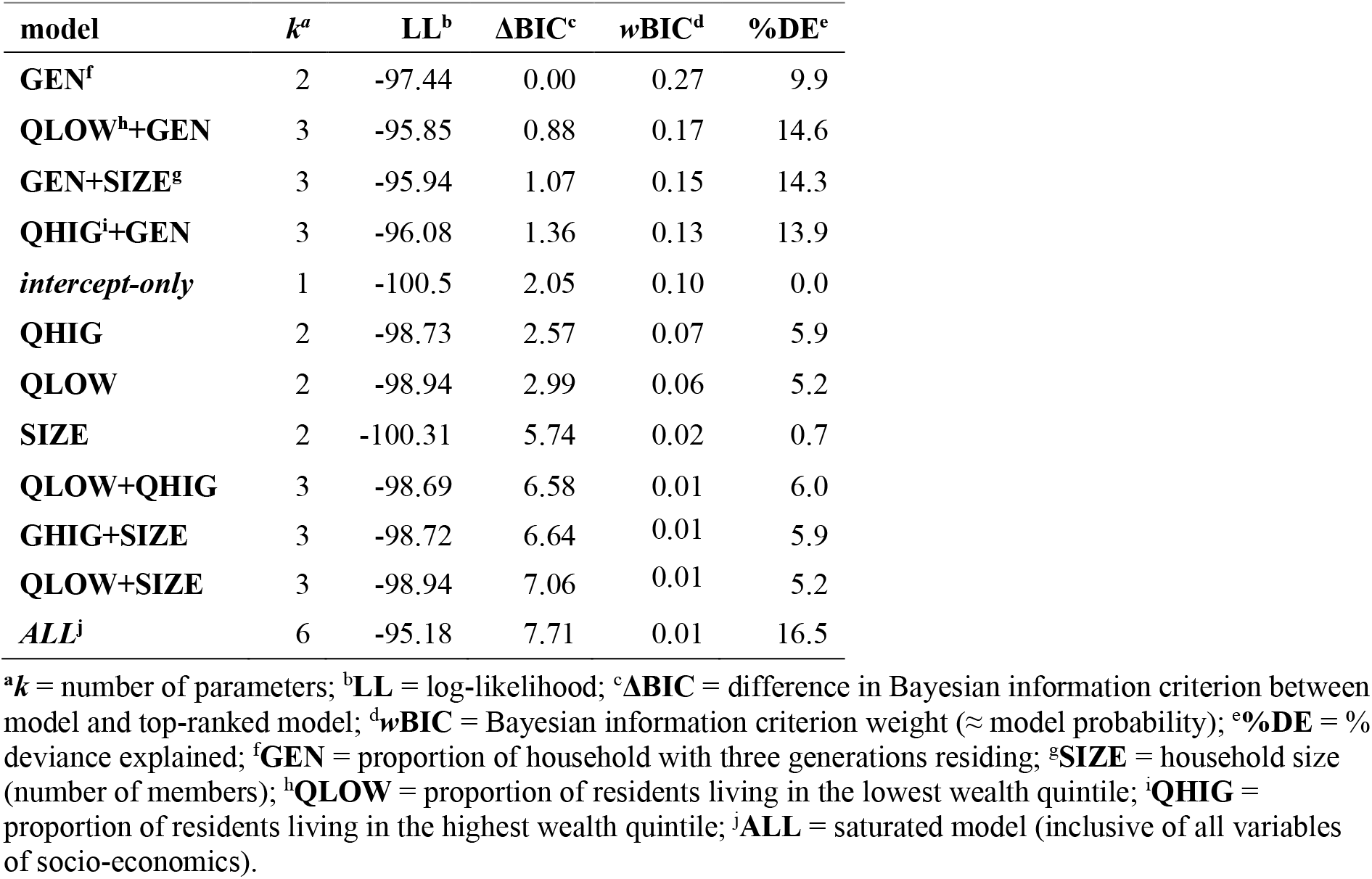
General linear models for indicators of socio-economics in relation to fertility among 59 low- and middle-income countries.

| model | $k^a$ | LL<sup>b</sup> | $\Delta BIC^c$ | $wBIC^d$ | %DE<sup>e</sup> |
| --- | --- | --- | --- | --- | --- |
| GEN<sup>f</sup> | 2 | -97.44 | 0.00 | 0.27 | 9.9 |
| QLOW<sup>h</sup>+GEN | 3 | -95.85 | 0.88 | 0.17 | 14.6 |
| GEN+SIZE<sup>g</sup> | 3 | -95.94 | 1.07 | 0.15 | 14.3 |
| QHIG<sup>i</sup>+GEN | 3 | -96.08 | 1.36 | 0.13 | 13.9 |
| *intercept-only* | 1 | -100.5 | 2.05 | 0.10 | 0.0 |
| QHIG | 2 | -98.73 | 2.57 | 0.07 | 5.9 |
| QLOW | 2 | -98.94 | 2.99 | 0.06 | 5.2 |
| SIZE | 2 | -100.31 | 5.74 | 0.02 | 0.7 |
| QLOW+QHIG | 3 | -98.69 | 6.58 | 0.01 | 6.0 |
| GHIG+SIZE | 3 | -98.72 | 6.64 | 0.01 | 5.9 |
| QLOW+SIZE | 3 | -98.94 | 7.06 | 0.01 | 5.2 |
| ALL<sup>j</sup> | 6 | -95.18 | 7.71 | 0.01 | 16.5 |

<sup>a</sup> $k$ = number of parameters; <sup>b</sup>LL = log-likelihood; <sup>c</sup> $\Delta BIC$ = difference in Bayesian information criterion between model and top-ranked model; <sup>d</sup> $wBIC$ = Bayesian information criterion weight ( ≈ model probability); <sup>e</sup>%DE = % deviance explained; <sup>f</sup>GEN = proportion of household with three generations residing; <sup>g</sup>SIZE = household size (number of members); <sup>h</sup>QLOW = proportion of residents living in the lowest wealth quintile; <sup>i</sup>QHIG = proportion of residents living in the highest wealth quintile; <sup>j</sup>ALL = saturated model (inclusive of all variables of socio-economics).

**Figure 5.**
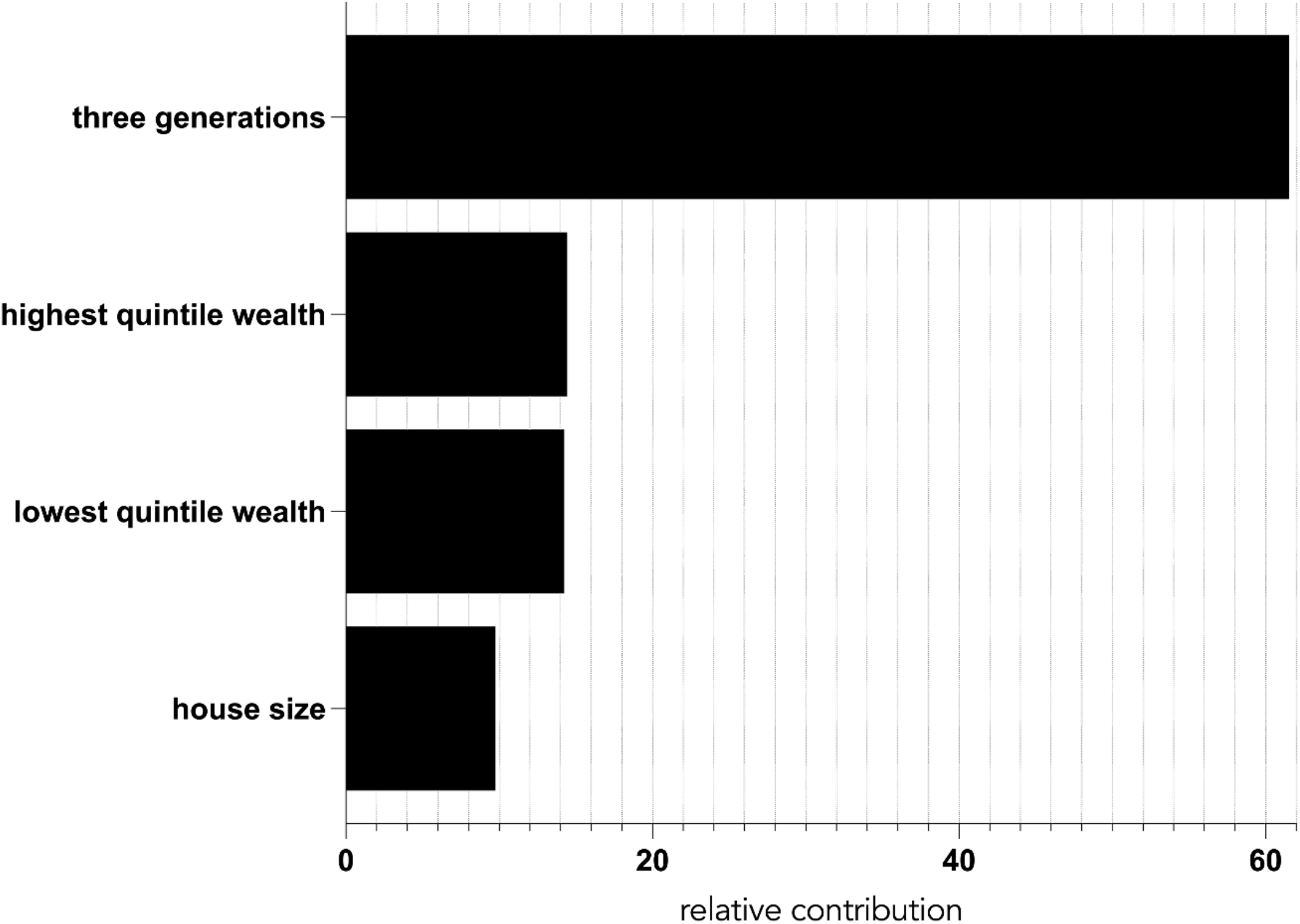
Boosted regression tree results (variable relative performance) for socio-economic indicators among 59 low- and middle-income countries. **three generations** = households with at least three generations residing; **house size** = mean number of household members; **highest quintile wealth** = proportion of a population residing in the highest wealth quintile (20%); **lowest quintile wealth** = proportion of a population residing in the lowest wealth quintile (20%).

